# Impact of Cushing’s syndrome on the hypothalamus-pituitary-gonad axis in women

**DOI:** 10.1101/2024.10.30.24316413

**Authors:** Anting Yu, Xuan Liu, Yiyu Chen, Shuo Li, Ming Liu

## Abstract

**Background:** The reproductive and sexual disorders commonly occur in patient with Cushing’s syndrome (CS), but only few clinical studies focused on the hypothalamus-pituitary-gonad (HPG) axis status in women with CS. A comprehensive spectrum of the impairment on HPG axis in women with CS of different tensity and causes is needed.

**Method:** This retrospective study analyzed the status of HPG axis in 137 women with different CS causes diagnosed between 2007 and May 2024, and the correlation between reproductive hormones and the tensity of hypercortisolism. Receiver operating characteristic (ROC) analysis was performed in 45 women with available data of plasma steroids by tandem mass spectrometry (LC-MS/MS) as well.

**Results:** Women with ectopic adrenocorticotropin (ACTH) secretion (EAS) had significantly higher levels of serum cortisol, 24h urinary-free cortisol (UFC), ACTH, with marked increase in testosterone and decrease in Luteinizing hormone (LH) and Follicle-stimulating hormone (FSH) (P<0.001).The serum cortisol and ACTH were positively associated with testosterone, while negatively associated with LH and FSH, especially in postmenopausal women. Further investigation of steroid profiles found plasma androgen including testosterone, Androstenedione (A2), dehydrospiandrostenedione (DHEA) and dehydrospiandrostenedione sulfate (DHEAS) had high sensitivity and specificity in discriminating CD from adrenal CS. Additional analysis of thyroid axis found hypercortisolism had less influence on TSH compared with LH and FSH.

**Conclusion:** Excessive cortisol caused by CS can impair the HPG axis in women, which were especially intense in EAS. The degrees of impairment were associated with the intensity and the underlying causes of hypercortisolism.

## Introduction

Cushing’s syndrome (CS), as a chronic and systemic endocrine disease caused by long-term and excessive exposure to glucocorticoids, can leads to a myriad of clinical manifestations and complications. CS traditionally comprises two subgroups: ACTH-dependent CS, which account for 70-80% of CS, and ACTH-independent CS (AI-CS).[1] ACTH-dependent CS is most commonly caused by Cushing’s disease (CD) (80-90%), and more rarely ectopic ACTH secretion (EAS) (10-20%), whose discrimination from Cushing’s disease is considered challenging[2, 3]. ACTH-independent CS is most frequently caused by unilateral adrenal adenomas (70-80%)[1]. The clinical complications of CS primarily include systematic arterial hypertension, impairment of glucose and lipid metabolism, musculoskeletal disorders, neuropsychiatric disorders, impairment of reproductive and sexual function and dermatological manifestations. Reproductive and sexual disorders commonly occur in patients with CS, and most frequently represented as decreased libido and hypogonadism in men and menstrual irregularity in women[4]. It is widely accepted that the hypothalamus-pituitary-gonad (HPG) axis can be affected by hypercortisolism at different levels, which accounts for the reproductive and sex disorders[5, 6]. Besides, excessive cortisol secretion can also impair the function of hypothalamus-pituitary-thyroid (HPT) axis[7].

Recently, some clinical studies has investigated the impact of CS on HPG axis in men, pointed out that hypercortisolism can lead to reversible hypogonadotropic hypogonadism in men[8, 9]. Though the female predominance exist in CS of different causes[1], only a few studies focused on the HPG status in women with CS in clinic[10–12], and rarely investigated the impairment of different subtypes of CS on HPG axis and its correlation with the tensity of hypercortisolism. Thus the aim of this retrospective study was to investigate the clinical status of HPG axis in women affected by different causes of CS and varying intensity of hypercortisolism.

## Materials and methods

### Setting

This was a retrospective study at the department of Endocrinology and Metabolism at Tianjin Medical University General Hospital. All coded medical records of women patient with CS assessed in routine care between 2007 and May 2024 were analyzed.

### Clinical evaluation

All patient were hospitalized at Tianjin Medical University General Hospital to undergo a full diagnostic work-up. The diagnosis of Cushing syndrome was based on clinical features and at least 2 of the following: (1) 24-h urinary free cortisol excretion (>300 nmol/24 h) in at least 2 samples; (2) 1-mg overnight dexamethasone suppression test (0800 h plasma cortisol>50 nmol/L) [13]; (3) 48 h, 2 mg/d dexamethasone suppression test urinary free cortisol>27 nmol/24 h [14]. Majority of the diagnoses of adrenal Cushing’s syndrome was histologically confirmed. The diagnosis of Cushing disease was established if pathological examination of the pituitary surgical specimen confirmed the diagnosis, or if the condition resolved after resection or irradiation of the pituitary gland. The diagnosis of an ectopic cause of ACTH secretion was confirmed by histological examination of surgical or biopsy specimen. Nonetheless, not all sources of ACTH excess were confirmed, in this case, presumed diagnosis was extracted from diagnostic code at discharge. The diagnoses were made at the discretion of the attending physician, based on clinical presentation, results of high-dose dexamethasone test, pituitary MRI, chest and abdominal CT.

### Hormone assays

Preoperative blood samples were obtained in the morning between 06:00 to 08:00 from an 8h fasting blood draw to minimize potential confounding introduced by circadian rhythmicity and food intake. Plasma ACTH, cortisol and 24-h excretion of urinary free cortisol were measured using chemiluminescent enzyme immunoassay (Siemens Healthcare Diagnostics Inc., Erlangen, Germany). The reproductive hormones (FSH, LH, estradiol, testosterone) were measured at the early time of primary diagnosis, and determined by chemiluminescence as well.

Only 45 women with CS were available to measure the plasma steroids by MS, and the steroid profile includes the following: progesterone, 17-hydroxyprogesterone, DHEA, DHEAS, androstenedione, testosterone, 11-deoxycorticosterone, 11-deoxycortisol, cortisol, corticosterone and cortisone. After appropriate sample preparation, the blood was centrifuged at 14,000g for 10 minutes. A 100 µL aliquot of the supernatant was then analyzed using a Jasper™ high-performance liquid chromatography system coupled with an ABSCIEX Triple Quad™ 4500 MD mass spectrometer, equipped with a heated nebulizer ionization source operating in positive ion mode. Quantification of the data was performed using MultiQuant™ MD 3.0.3 software.

### Statistic analysis

Statistical analysis was performed using GraphPad Prism version 9.0, IBM SPSS Statistics version 25 and R (a language and enviroment for statistical computing). Standard descriptive analyses were used to summarize the study variables. Shapiro’s test was applied to test for normality. For quantitative variables,comparisons between controls and cases at baseline were performed by unpaired Student’s t-tests (or Mann-Whitney tests if the distribution of the variable was not log-normal). Comparison between the three subgroups of CS was conducted by ordinary one-way ANOVA in case of homoscedasticity or Kruskal-Wallis instead. The simple linear regression was used to study the association between Cushing’s indexes and reproductive outcomes at baseline. The receiver operating characteristic (ROC) curve analysis was performed to test the usefulness of plasma androgens as markers for differentiating between adrenal CS and CD.The optimal cutoff values were determined using the Youden index (sensitivity + specificity −1). A p-value <0.05 was considered statistically significant.

## Results

### Demographic and clinicopathological characteristics

287 female patients clinically suspected as Cushing’s syndrome were considered for inclusion in this study to begin with. After the exclusion of subclinical CS, iatrogenic CS and patients lacking sexual hormone assays, only 137 women with CS served as study’s population, including 63 with adrenal CS, 57 with CD and 17 with EAS. Of note, the source of ectopic ACTH secretion include 5 thymic neuroendocrine tumor, 3 gastrointestinal carcinomas, 2 pheochromocytomas, 1 renal clear cell carcinoma, 1 lung neuroendocrine tumor, 1 small cell lung carcinoma and 1 pancreatic neuroendocrine tumor.

### Gonadotrope axis in women patient with Cushing’s syndrome

The basic information and hormonal characteristics of study’s cohort are shown in Table 1. No significant difference in body mass index (BMI) were showed. But the mean age of EAS was older than the other two subgroups, comport with previous report [3]. The three subgroups of women with CS have markedly different levels of ACTH (AI-CS<CD<EAS), whereas the intensity of hypercortisolism, represented by serum cortisol and 24h-UFC, only significantly higher in women with EAS (P<0.001).

**Table 1.** Characteristics of 137 women with Cushing’s syndrome.

|  | **Adrenal Cushing's**<br>**syndrome**<br>**(n=63)** | **Cushing's**<br>**disease**<br>**(n=57)** | **ectopic ACTH**<br>**syndrome**<br>**(n=17)** | **P-value** |
| --- | --- | --- | --- | --- |
| Age<br>(years) | 40.11±12.60 | 39.95±11.09 | 55.65±16.15 | **<0.001** |
| Body mass index<br>(BMI)(kg/m<sup>2</sup>) | 26.15±4.19 | 26.92±4.83 | 27.09±5.31 | 0.587 |
| ACTH<br>(pg/ml) | 5.00(5.00,6.79) | 66.40(46.55,114.00<br>) | 225.00(123.00,481.<br>00) | **<0.001** |
| Serum cortisol<br>(ng/ml) | 30.85±8.75 | 33.07±11.53 | 51.04±17.01 | **<0.001** |
| Urinary-free cortisol<br>(mg/24 h) | 450.00(193.50,650.<br>00) | 412.20(174.65,592.<br>80) | 1424.50(945.00,208<br>1.00) | **<0.001** |
| FSH<br>(IU/L) | 4.54(3.00,7.48) | 5.18(3.58,8.25) | 1.70(0.79,3.32) | **<0.001** |
| LH<br>(IU/L) | 3.29(1.32,6.21) | 2.58(1.06,6.92) | 0.16(0.07,0.39) | **<0.001** |
| E2<br>(pg/ml) | 34.00(15.00,91.00) | 32.86(14.71,54.00) | 22.80(14.50,31.67) | 0.365 |
| T<br>(ng/dl) | 24.15(17.79,40.23) | 52.00(38.77,72.78) | 105.50(65.26 ,<br>199.80) | **<0.001** |
| FSH:LH ratio | 2.01(1.20,3.87) | 2.62(1.64,4.64) | 10.83(3.41,20.78) | **<0.001** |
| FT3<br>(pmol/L) | 3.21±0.66 | 3.29±0.81 | 2.64±0.72 | **0.005** |
| FT4<br>(pmol/L) | 11.274±1.778 | 11.556±2.470 | 11.86±3.54 | 0.613 |
| TSH<br>(uIU/mL) | 0.69(0.53,1.22) | 0.62(0.36,1.14) | 0.49(0.23,0.91) | 0.097 |
| FT3:FT4 ratio | 0.29(0.25,0.31) | 0.28(0.25,0.32) | 0.23(0.18,0.27) | **<0.001** |

Statistically significant results(P<0.05) were highlighted in bold.Comparison between groups was performed using one-way ANOVA or Kruskal-Wallis test if the distribution of the respective variable was not log-normal in any of the groups. ACTH, adrenocorticotrophic hormone; FSH, Follicle-stimulating hormone; LH, Luteinizing hormone; E2, estradiol; T, testosterone; TSH, thyroid-stimulating hormone.

Comparisons of reproductive hormones between women with different CS are detailed in Figure1. There was no much difference in serum estradiol level between adrenal CS, CD and EAS. Conversely, the testosterone levels significantly increased in women with EAS and CD in comparison with AI-CS (P<0.001), and rather higher in EAS (P<0.001) (Figure 1 A, B). Both circulating LH and FSH levels in women with EAS markedly decreased, showing significant differences with the other two subgroups. But the LH and FSH levels of women with CD did not significantly differ from AI-CS (Figure 1 C, D). Furthermore, the FSH:LH ratios were also calculated and compared, and the EAS showed markedly higher FSH:LH ratio than AI-CS (P<0.001) and CD (P=0.002), while no marked difference existed between AI-CS and CD (supplementary figure S1).

**Figure 1.**
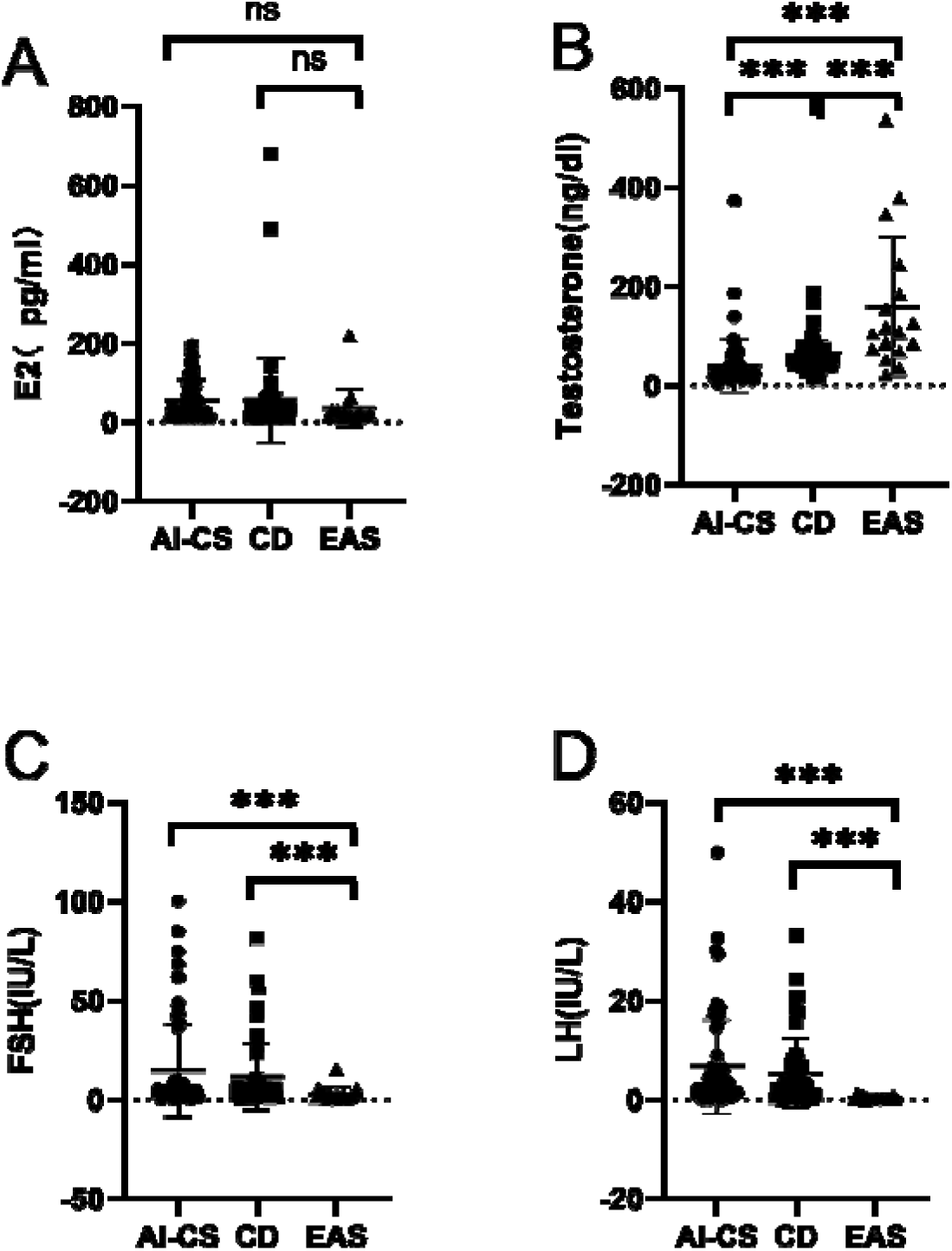
Scatter plots of reproductive hormones. Estradiol, testosterone, LH and FSH values are displayed in (A)-(C), respectively. All comparison between the three subgroups are shown by brackets, asterisks or text. E2, estradiol; FSH, Follicle-stimulating hormone; LH, Luteinizing hormone; AI-CS, ACTH-independent Cushing’s syndrome; CD, Cushing’s disease; EAS, ectopic ACTH syndrome. ***, P<0.001; ns, not significant.

### Linear regression between hypercortisolism intensity and reproductive hormones in premenopausal and postmenopausal women

It is a general consensus that the level of women’s reproductive hormones have clear periodicity and regulate the menstruation [15], thus the study’s group was further divided into the premenopausal women and the postmenopausal women, detailed in Table 2. As shown, most women with EAS were postmenopausal (premenopause, n=5; psomenopause, n=12), with significantly higher levels of serum cortisol, 24h-UFC, ACTH and testosterone levels, and lower levels of LH and FSH. It is noteworthy that the LH and FSH levels reached a more significant difference in postmenopausal women with EAS in comparison with adrenal CS and CD (Supplementary Figure S2). The serum cortisol and 24h UFC levels did no significantly differ between AI-CS and CD neither in premenopause nor postmenopause women, while the ACTH and testosterone levels were higher in CD than AI-CS in both situations, similar with above (Table 1). The estradiol levels still showed no difference among the three subgroups neither in premenopausal women nor in postmenopausal women.

**Table 2.**
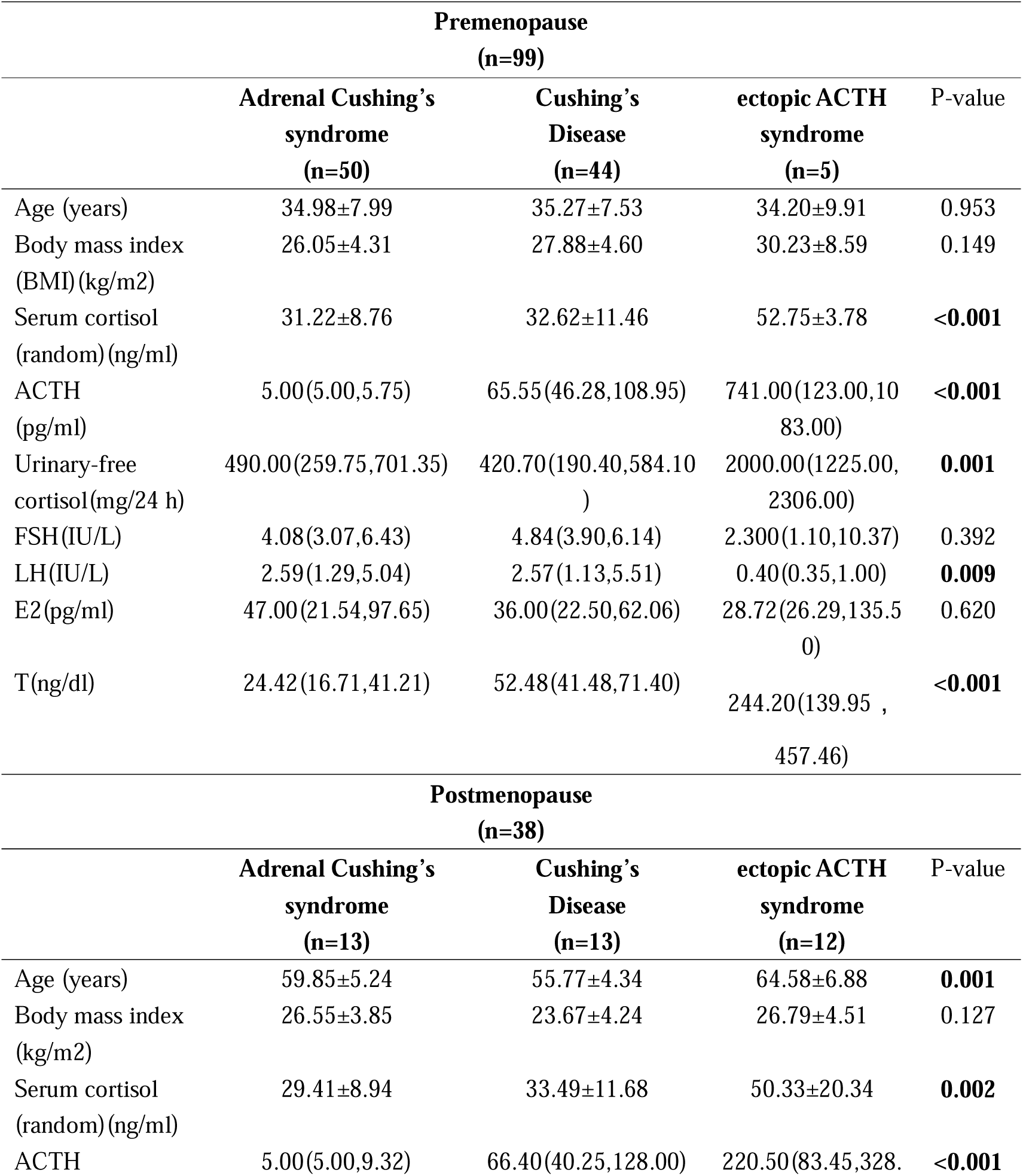
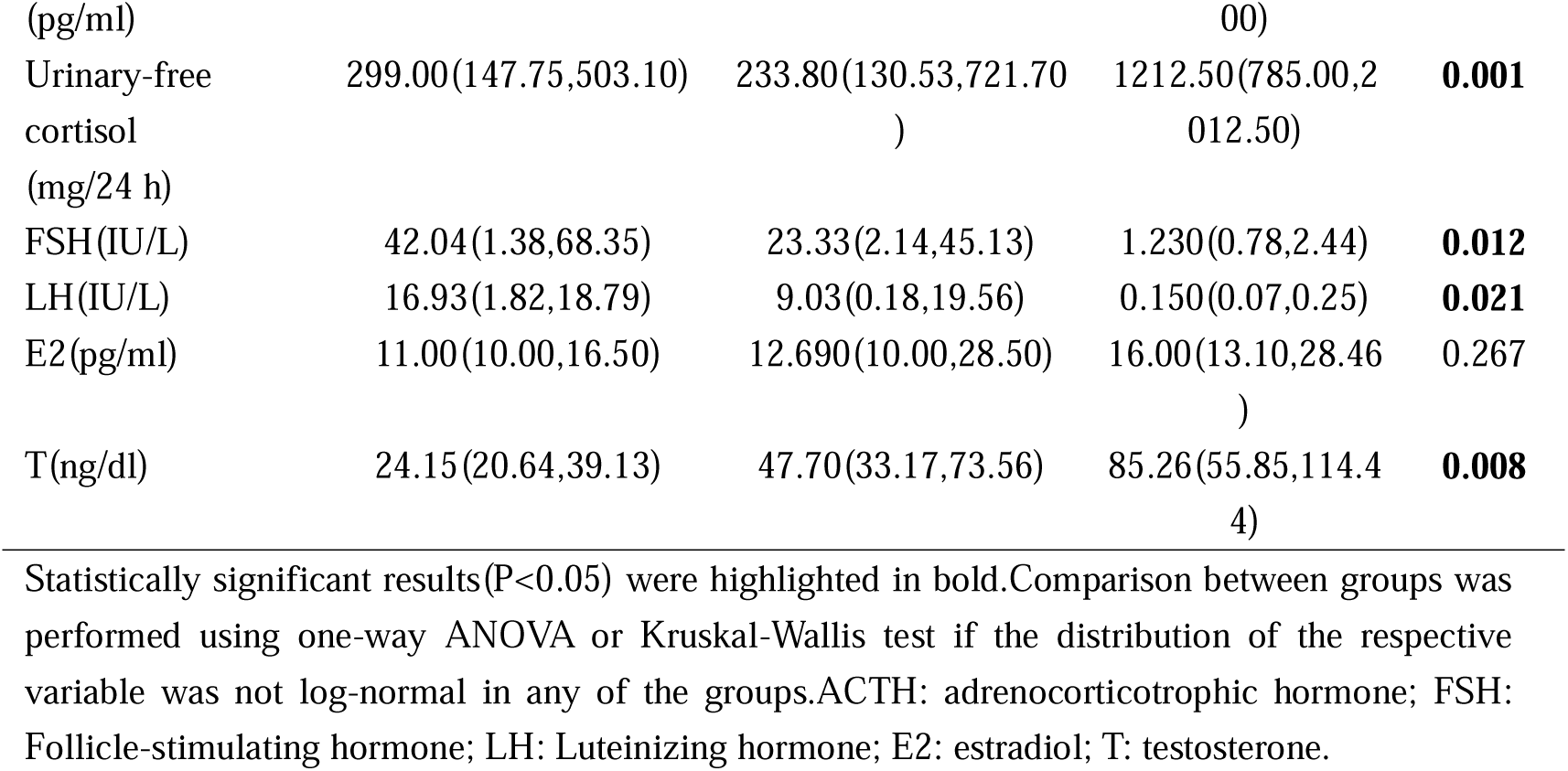
Characteristics of women with Cushing’s syndrome separated by menstruation.

For the sake of investigating the relationship between the tensity of hypercortisolism and the HPG axis impairment, linear regression analyses between serum cortisol and the reproductive hormones were performed, detailed in Figure 2. As shown, in postmenopausal women, serum cortisol was significantly and negatively associated with FSH (P=0.014) and LH (P=0.016), while positively associated with testosterone (P<0.001). Similar analyses were performed in premenopausal women with CS as well, and found that serum cortisol did not significantly associated with LH or FSH. Only testosterone was positively associated with serum cortisol in premenopausal women with CS (P<0.001). Of note, the linear regression curves in postmenopausal women described those relationships better, which are demonstrated by Pearson’s correlations[16].

**Figure 2.**
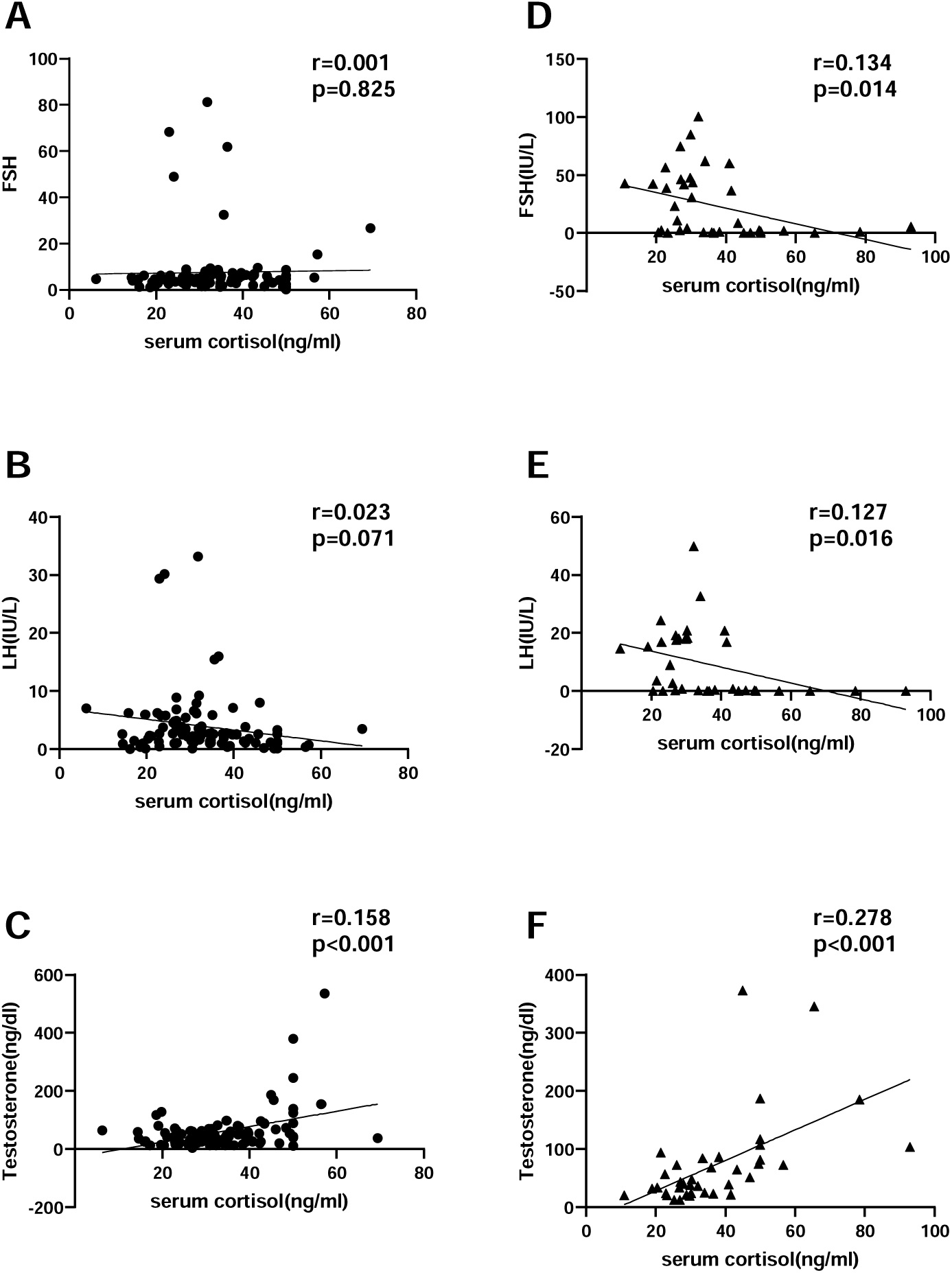
Linear regressions between serum cortisol and reproductive hormones in premenopausal and postmenopausal women. Premenopausal women with Cushing’s syndrome were included in (A)-(C). Postmenopausal women with CS were included in (D)-(F) to avoid effects of the menstrual cycle. Pearson correlations(r) were calculated squares. FSH, Follicle-stimulating hormone; LH, Luteinizing hormone.

The linear regression between ACTH, 24h-UFC and reproductive hormones are presented in Supplementary Table S1. The results were quiet similar with serum cortisol when studying the relationship between ACTH and reproductive hormones, showing significant correlations with testosterone in both situations, but only in postmenopausal women ACTH did significantly correlate with LH and FSH. Moreover, the 24h-UFC only showed significant correlation with testosterone, and merely did correlate with LH or FSH in neither situations. Still, serum estradiol did not correlate with any parameters of CS in any situations.

### The characteristics of steroid profiles in adrenal CS and Cushing’s disease

The measurement of plasma steroids by MS was only available in 45 women with CS, detailed in Table 3 (AI-CS, n=24; CD, n=21). As shown, plasma androgen including testosterone, A2, DHEA and DHEAS significantly increased in women with CD compared with AI-CD (P<0.001) (Supplementary Figure S3). Additional linear regression analyses between ACTH and DHEA and DHEAS were performed as well, and showed significant and positive association between them (supplementary figure S4), comport with the general acceptance that DHEA and DHEAS are majorly determined by ACTH levels [17]. Except plasma androgen, only 11-Deoxycortisol (P=0.011) and 11-deoxycortisterone (P=0.006) moderately decreased in women with CD.

**Table 3.**
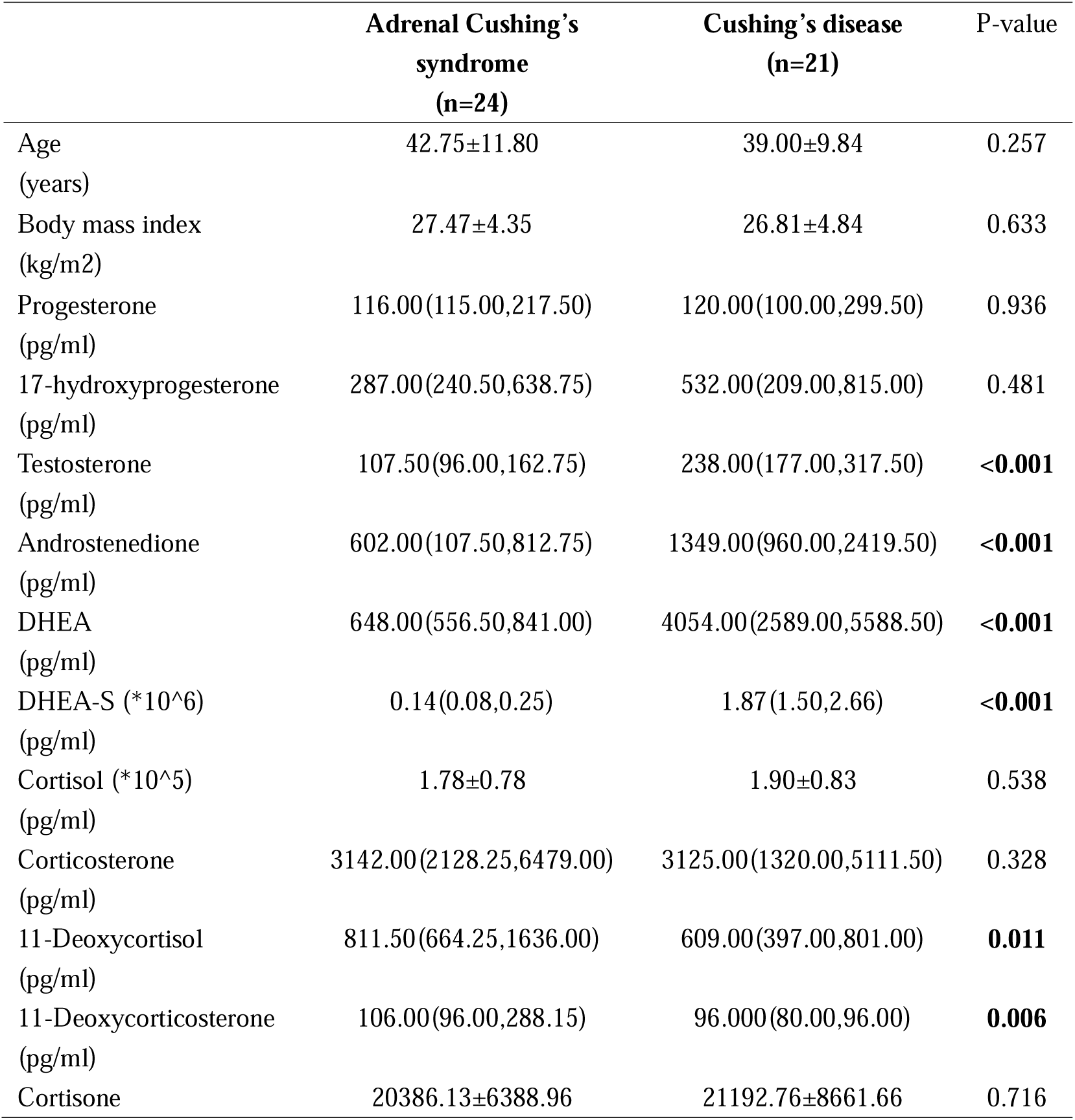
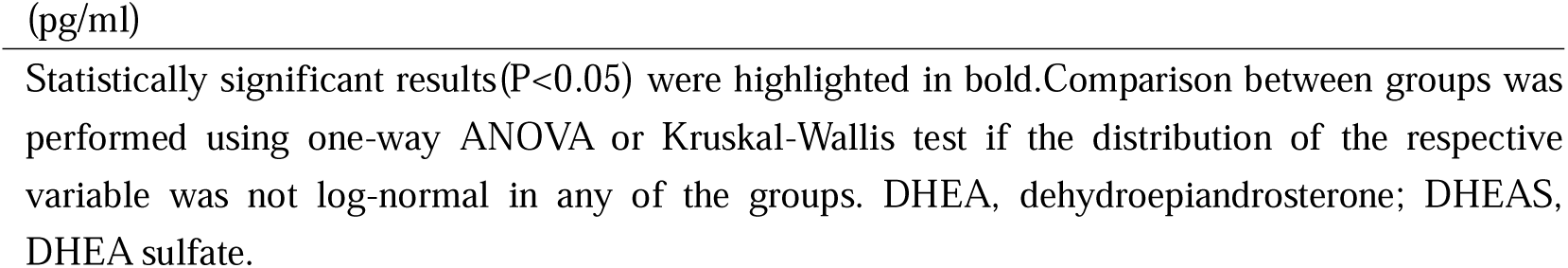
Characteristics of steroid profile in 45 women with CS.

|  | Adrenal Cushing's syndrome (n=24) | Cushing's disease (n=21) | P-value |
| --- | --- | --- | --- |
| Age (years) | 42.75±11.80 | 39.00±9.84 | 0.257 |
| Body mass index (kg/m2) | 27.47±4.35 | 26.81±4.84 | 0.633 |
| Progesterone (pg/ml) | 116.00(115.00,217.50) | 120.00(100.00,299.50) | 0.936 |
| 17-hydroxyprogesterone (pg/ml) | 287.00(240.50,638.75) | 532.00(209.00,815.00) | 0.481 |
| Testosterone (pg/ml) | 107.50(96.00,162.75) | 238.00(177.00,317.50) | **<0.001** |
| Androstenedione (pg/ml) | 602.00(107.50,812.75) | 1349.00(960.00,2419.50) | **<0.001** |
| DHEA (pg/ml) | 648.00(556.50,841.00) | 4054.00(2589.00,5588.50) | **<0.001** |
| DHEA-S (*10^6) (pg/ml) | 0.14(0.08,0.25) | 1.87(1.50,2.66) | **<0.001** |
| Cortisol (*10^5) (pg/ml) | 1.78±0.78 | 1.90±0.83 | 0.538 |
| Corticosterone (pg/ml) | 3142.00(2128.25,6479.00) | 3125.00(1320.00,5111.50) | 0.328 |
| 11-Deoxycortisol (pg/ml) | 811.50(664.25,1636.00) | 609.00(397.00,801.00) | **0.011** |
| 11-Deoxycorticosterone (pg/ml) | 106.00(96.00,288.15) | 96.000(80.00,96.00) | **0.006** |
| Cortisone | 20386.13±6388.96 | 21192.76±8661.66 | 0.716 |

Statistically significant results(P<0.05) were highlighted in bold.Comparison between groups was performed using one-way ANOVA or Kruskal-Wallis test if the distribution of the respective variable was not log-normal in any of the groups. DHEA, dehydroepiandrosterone; DHEAS, DHEA sulfate.

The receiver operating characteristic curve (ROC) analyses were applied to assess the diagnostic power of plasma androgen in differentiating CD from adrenal Cushing’s syndrome, and detailed in Table 4 and Figure 3. As shown, the testosterone had a sensitivity of 81.0% and a specificity of 83.3%, with a cutoff value of 172.5 pg/ml and an AUC of 0.816. The androstenedione had a sensitivity of 100.0% and a specificity of 83.3%, with a cutoff value of 917.5 pg/ml and an AUC of 0.829. The DHEA had a sensitivity of 100.0% and a specificity of 87.5%, with a cutoff value of 1170.5 pg/ml and an AUC of 0.972. The DHEA-S had a sensitivity of 100.0% and a specificity of 83.3%, with a cutoff value of 0.27*10^6 pg/ml and an AUC of 0.958. Taken together, all four plasma androgen can effectively discriminate between adrenal CS and CD, with high sensitivity and specificity at the best cutoff, especially DHEA and DHEAS.

**Figure 3.**
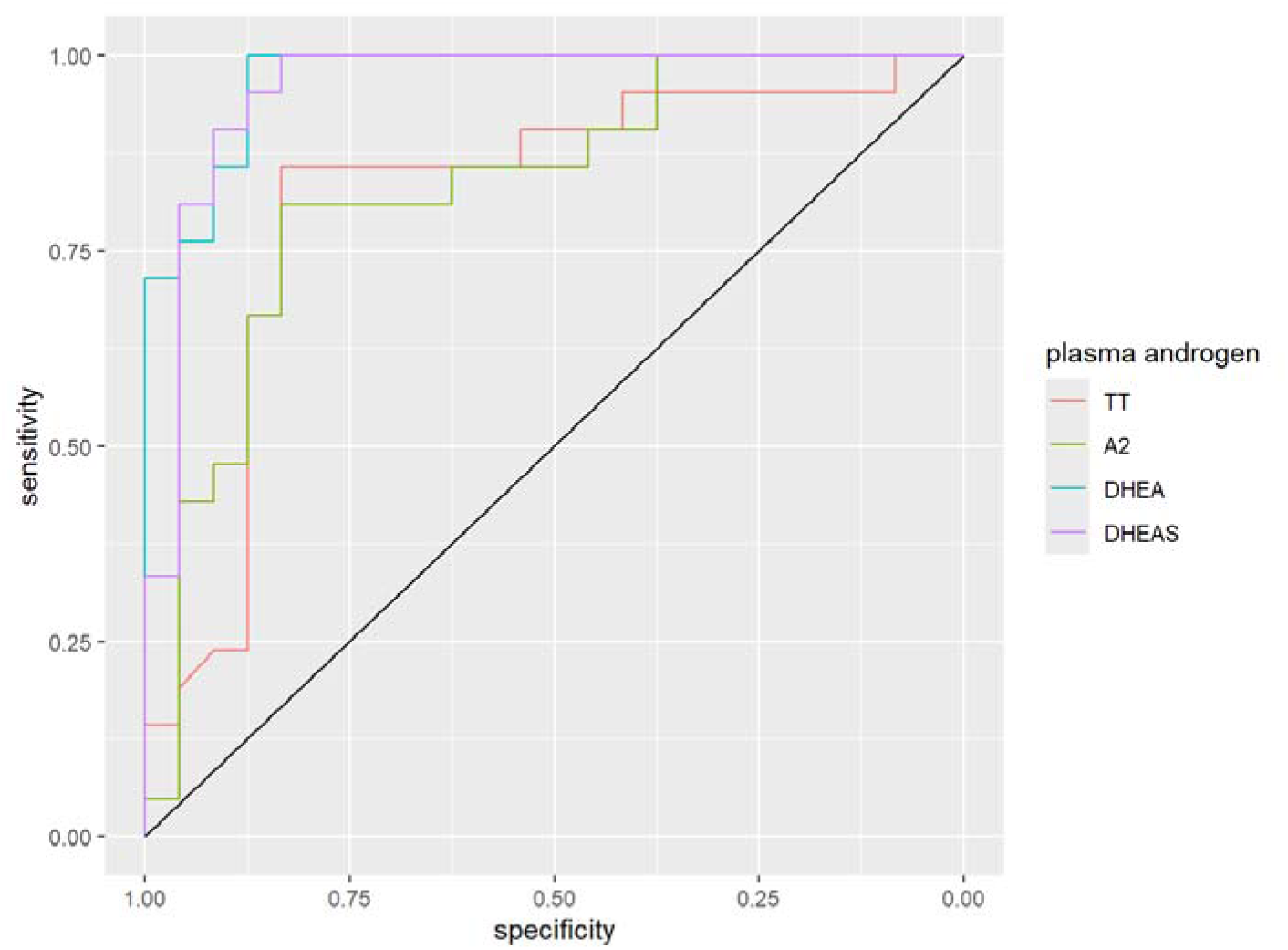
ROC-analysis to different between adrenal CS and CD with plasma androgens. ROC-analysis to differentiate between patients with AI-CS and CD with the most discriminative steroids. TT, total testosterone; A2, androstenedione; DHEA, dehydrospiandrostenedione; DHEAS, dehydrospiandrostenedione sulfate.

**Table 4.**
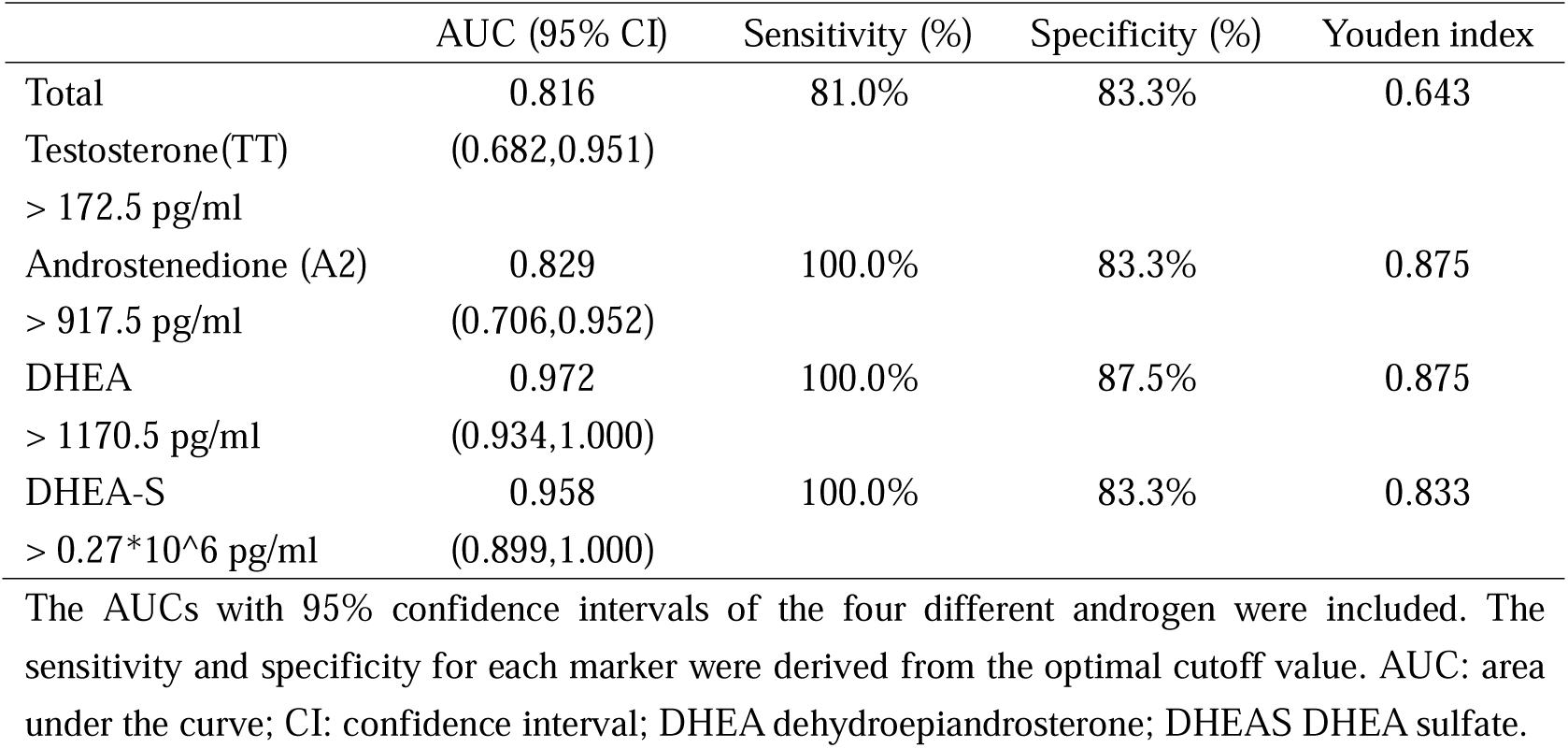
Accuracy of steroid biomarkers for differentiating CD form adrenal CS.

### Thyroid axis in women patient with Cushing’s syndrome

Due to the Interest in the impact of hypercortisolism on HPT axis, and the high prevalence of thyroid function test, the data of thyroid hormone was also collected, detailed in Table1 and Figure 4. As shown, only serum FT3 levels in women with EAS markedly decreased compared to adrenal CS and CD (P<0.01), either FT4 nor TSH showed much difference (figure 5A, B). The FT3:FT4 ratios were further calculated and compared, and reflected a more distinguishing difference in EAS compared with AI-CS and CD as well(P<0.001) (Figure 5C), which statistically indicated the prominent effect of cortisol-meditated inhibition on peripheral deiodination [7]. The linear regression analyses between hypercortisolism and thyroid hormones were also performed, detailed in supplementary figure S5. Only the FT3 showed a moderate correlation with serum cortisol, 24h-UFC and ACTH, neither FT4 nor TSH did significantly correlate with any parameters of CS.

**Figure 4.**
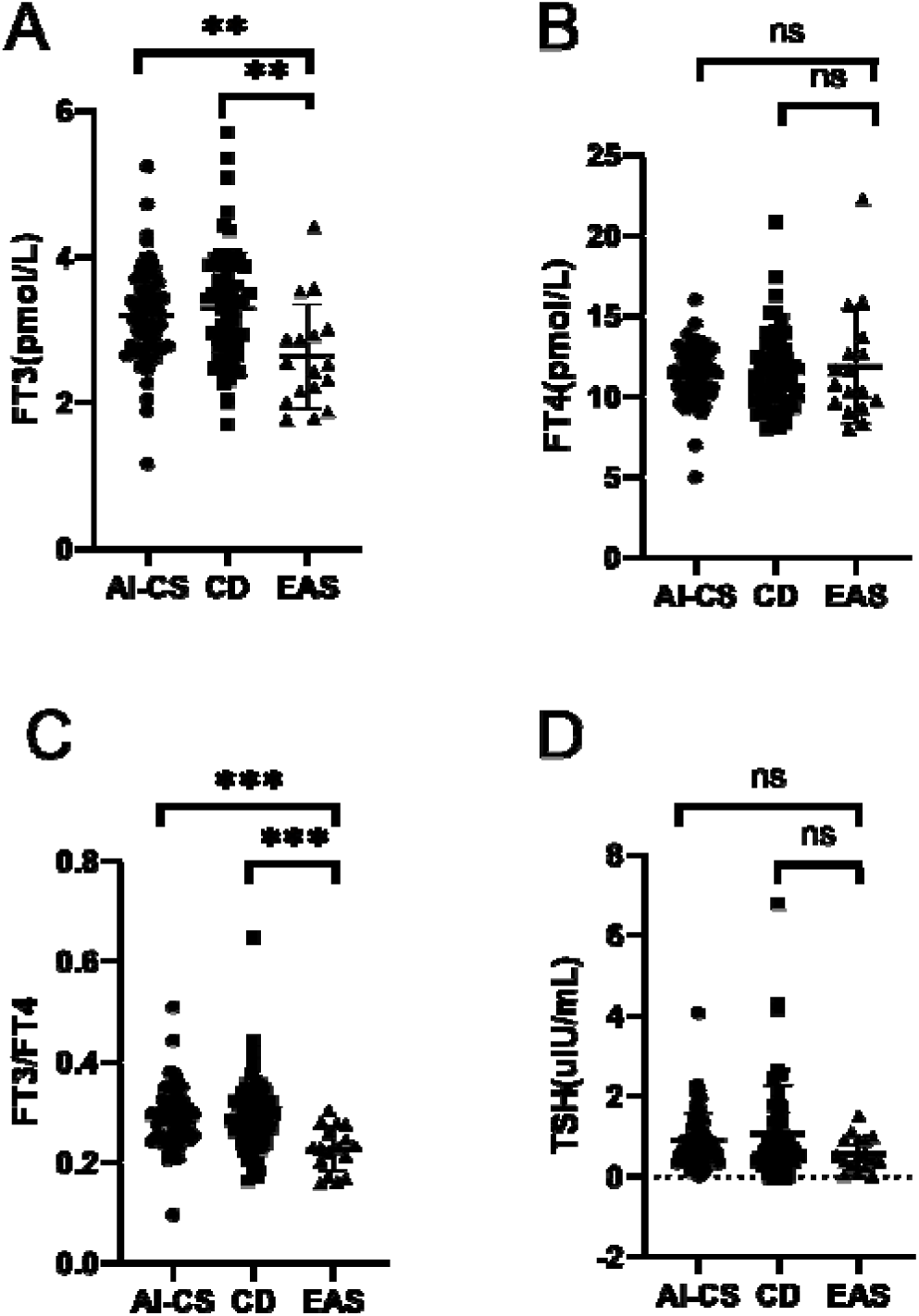
Scatter plots of Thyroid hormone. Serum FT3,FT4, TSH and FT3/FT4 values are displayed in(A)-(D).All comparison between the three subgroups are shown by brackets,asterisks or text. TSH: thyroid-stimulating hormone; AI-CS, ACTH-independent Cushing’s syndrome; CD, Cushing’s disease; EAS, ectopic ACTH syndrome. ***, P<0.001; **, P<0.01; *, P<0.05; ns, not significant.

## Discussion

This study successfully characterizes the impact of hyercortisolism on gonadotrope axis in women with CS of different causes, and further clarifies its correlation with the intensity of hypercortisolism. The steroids profiles between CD and adrenal CS in a limited group is investigated as well, offering a more profound insight into the changes of sexual hormones. The effects of CS on HPT axis is also mentioned, in comparison with the HPG axis.

The primary finding of this study is the distinguishing decrease of LH and FSH, and the marked increase of testosterone levels in women with EAS compared with adrenal CS and CD. It is wildly accepted that EAS is usually associated with more severe hypercortisolism, which was also confirmed in this study, and that glucocorticoids can dose-dependently impair the hypothalamus, pituitary and the reproductive tract in women[1, 5, 6], thus the decrease of LH and FSH is theoretically expected. However, there was no significant difference in estradiol between the three CS subgroups, nor correlation with the tensity of hypercortisolism in neither premenopausal cohort or in the postmenopausal cohort. This may be explained by the dual roles of glucocorticoids in the ovary. The glucocorticoids have been found have both stimulatory and inhibitory effects on ovary, depending on the cortisol dosage, follicular phase or GR isoforms expression, and the function still remains unclear [18]. Moreover, women with EAS had markedly higher FSH:LH ratio, and there has study pointed out that a basal cycle high FSH:LH ratio has a negative impact on follicular development and oocyte quality [19], indicating another possible mechanism by which the gonad was impaired in women with EAS.

This study also found significant linear associations between serum cortisol and ACTH versus reproductive hormones, especially in postmenopausal women with CS, which were negative in LH and FSH, while positive in testosterone. Through further dividing the study’s group by the menopause, we hope to exclude the influence of menstruation on reproductive hormones. And the result exhibited more fitting models of linear regression for the association between the tensity of hypercortisolism and LH and FSH in postmenopausal cohort, demonstrated by higher Pearson’s correlations [16]. Comport with previous studies [5, 6, 18], those results once again confirmed that the function of HPG axis in women can be impaired by hypercortisolism, and is associated with its intensity. Nevertheless, similar with previous study in men, these associations were quite moderate[8], indicating that other influential factors, like personal heterogeneity and underlying cause, also have influence on the degrees of gonadal impairment by hypercortisolism.

As mentioned above, the response of testosterone to hypercortisolism was antithetical to LH and FSH, showing that more severe hypercortisolism is associated with higher plasma testosterone in women with CS. Further, the steroids profile by MS showed that testosterone, A2, DHEA and DHEAS were all increased in women with CD compared with AI-CS. Interestingly, this finding is right inverse to the finding in men, which showed that testosterone levels were frequently lower in men with CS and negatively associated with the tensity of hypercortisolism [8, 9, 20]. This phenomenon suggests that the sexual dimorphism exists in testosterone’s response to CS. The explanation lies in that the testosterone is mainly secreted by testicle in men, and its secretion can be inhibited by hypercortisolism [20], whereas in women, the testosterone is produced from adrenal (25%), ovary (25%), and circulating A2 (50%), and the A2 is secreted by adrenal (50%) and ovary (50%) as well [21]. As generally accepted, the adrenal androgen production is stimulated by ACTH [22], which is also confirmed by this study’s result, presenting significant and positive correlation between ACTH and plasma androgen. Thus in women with CS, the regulation of ACTH on adrenal androgen seemed have a more predominant role than the inhibition of hypercortisolism on gonad.

The steroid profiles of CS based on MS is a hot topic recently, many studies have reported that the steroid profiles can help diagnose different subtypes of CS, including clinical and subclinical CS, adrenal CS, CD, EAS and so on [23–26]. The diagnostic power of plasma androgen in discriminating CD and adrenal CS was also analyzed in this study, and found all the four androgen can effectively differentiate CD from adrenal CS. Among them, DHEA and DHEAS can best serve as diagnostic parameters with higher sensitivity and specificity, making them useful supplementary measurement to confirm the causes of CS.

This study also investigated the impact of CS on HPT axis, but only found the peripheral inhibitory effect of hypercortisolism, demonstrated by the decreased FT3 and T3:T4 ratio in EAS, and the negative correlation between FT3 and hypercortisolism. The FT4 and TSH showed no significant difference or correlation with hypercortisolism, which seemed contrary to previous study [7, 27]. But this result may imply that the HPG axis was more vulnerable to excessive serum cortisol than the thyroid axis, since the HPG axis seemed more severely impaired than thyroid axis but similar tensity of hypercortisolism. As normal thyroid function is important to maintaining normal reproduction, and thyrotoxicosis and hypothyroidism can cause reproductive and sex disorders in both men and women [28], extra attentions should be paid on HPG axis in those patient with CS and hypothyroidism.

The major limitation of this study is its retrospective design, which inevitably leading to selection bias, personal heterogeneity and measurement incompleteness. Besides, the 24h-UFC merely had significant correlations with reproductive hormones in this study, which may attributed to the retrospective design and the high degree of variability in UFC levels [29].The limited number available to MS is also a limitation, due to the low prevalence of MS, but taken previous studies together [23, 24, 30], the conclusion about the steroid profiles is quiet convincing.

The major strength of this study is that it met the need to characterize the clinical spectrum of CS on HPG axis in women with the large population. Though it is wildly received that high dose of glucocorticoid can impair reproductive system [5, 6, 18], only a handful studies reported its clinical status[10–12]. This study further divided the whole cohort by etiology and menopause, and through investigating the differences and associations among different groups, offered more comprehensive insight into the influence of hypercortisolism on HPG axis. The involvement of steroid profile using MS is also the novelty of this study, provided support for plasma androgen in discriminating CD from adrenal CS. Moreover, with the wild use of glucocorticoids, the result of this study highlights the importance to concern the HPG axis status in women with exogenous CS as well, especially in those who seek fertility.

In conclusion, the impairment of Cushing’s syndrome on hypothalamic-pituitary-gonadal axis in women is associated with the underlying causes and the intensity of hypercortisolism. Women with ectopic ACTH syndrome incline to show a more severe impairment of HPG axis but higher testosterone levels.

## Data Availability

All data produced in the present study are available upon reasonable request to the authors.

## Acknowledgments

We thank all participants from the Department of Endocrinology and Metabolism at Tianjin Medical University General Hospital.

## Ethics approval

The studies involving human participants were reviewed and approved by Institutional Review Board of Tianjin Medical University General Hospital. The ethics committee waived the requirement for written informed consent for participation.

## Human participant declaration

Due to the extraction of all patient data from the hospital’s electronic medical records and the anonymization of participants’ identities, the study waived the requirement for informed consent.

## Statement of authorship

Anting Yu participated in data collection and drafting and editing of the paper.

Ruixuan Liu, Yiyu Chen and Shuo Li participated in data analysis.

Ming Liu participated in revision of the paper.

All authors saw and approved the final version and no other person made a substantial contribution to the paper.

