## Supplementary Figure S1 for "Impact of Cushing’s syndrome on the hypothalamus-pituitary-gonad axis in women"

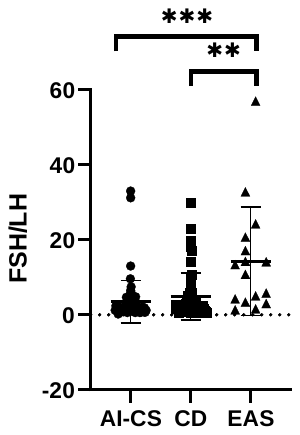

**Supplementary Figure S1. Scatter plots of FSH:LH ratios.** All comparison between the three subgroups are shown by brackets,asterisks or text. FSH, Follicle-stimulating hormone; LH, Luteinizing hormone; AI-CS, ACTH-independent Cushing’s syndrome; CD, Cushing’s disease; EAS, ectopic ACTH syndrome. ***, P<0.001; **, P<0.01.
