## Supplementary Figure S2 for "Impact of Cushing’s syndrome on the hypothalamus-pituitary-gonad axis in women"

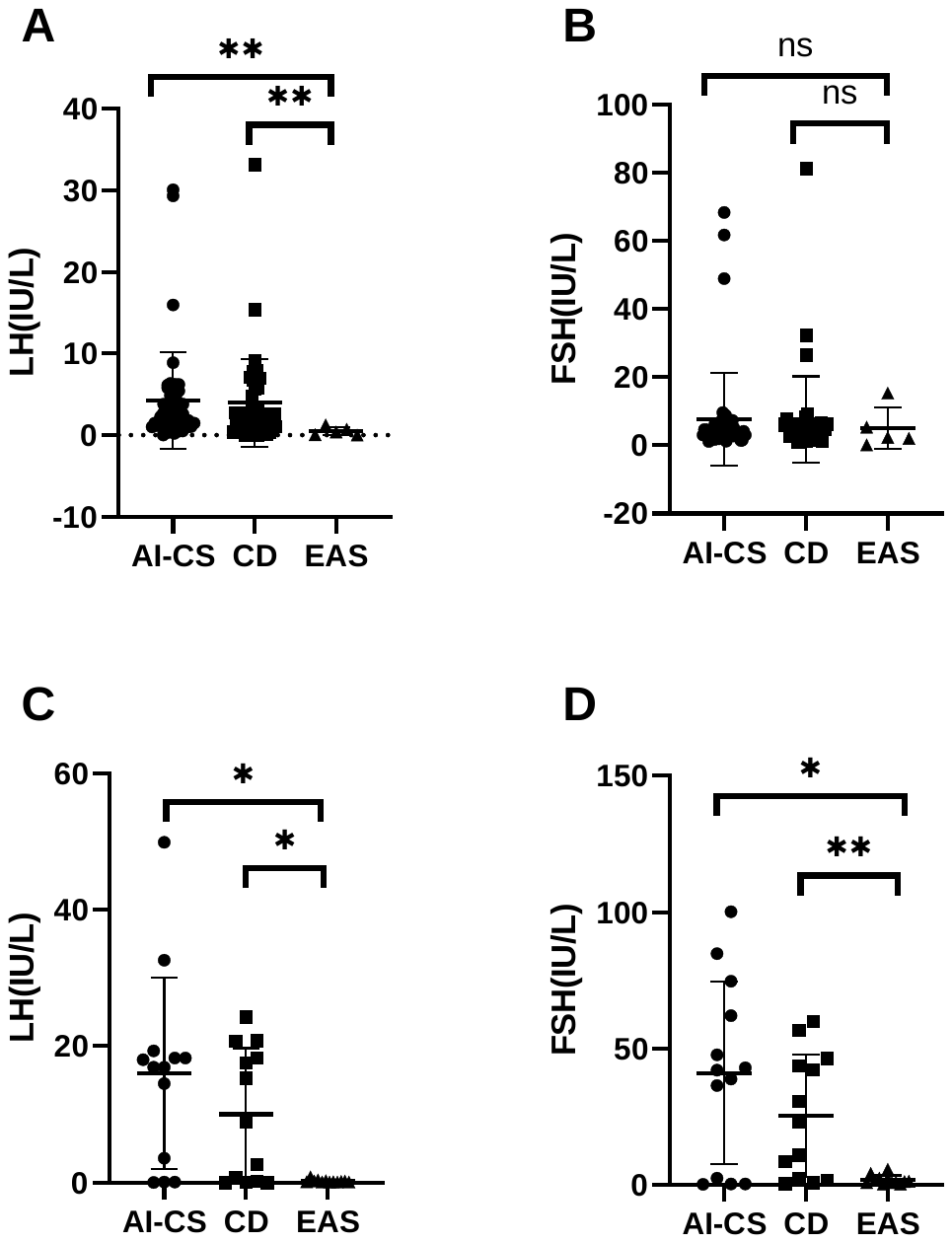

**Supplementary Figure S2. Scatter plots of LH and FSH in women separated by menstruation.** (A)-(B), premenopausal women with CS were included. (C)-(D), postmenopausal women with CS were included. All comparison are shown by brackets,asterisks or text. FSH, Follicle-stimulating hormone; LH, Luteinizing hormone; AI-CS, ACTH-independent Cushing’s syndrome; CD, Cushing’s disease; EAS, ectopic ACTH syndrome. ***, P<0.001; **, P<0.01; *, P<0.05; ns, not significant.
