## Supplementary Figure S3 for "Impact of Cushing’s syndrome on the hypothalamus-pituitary-gonad axis in women"

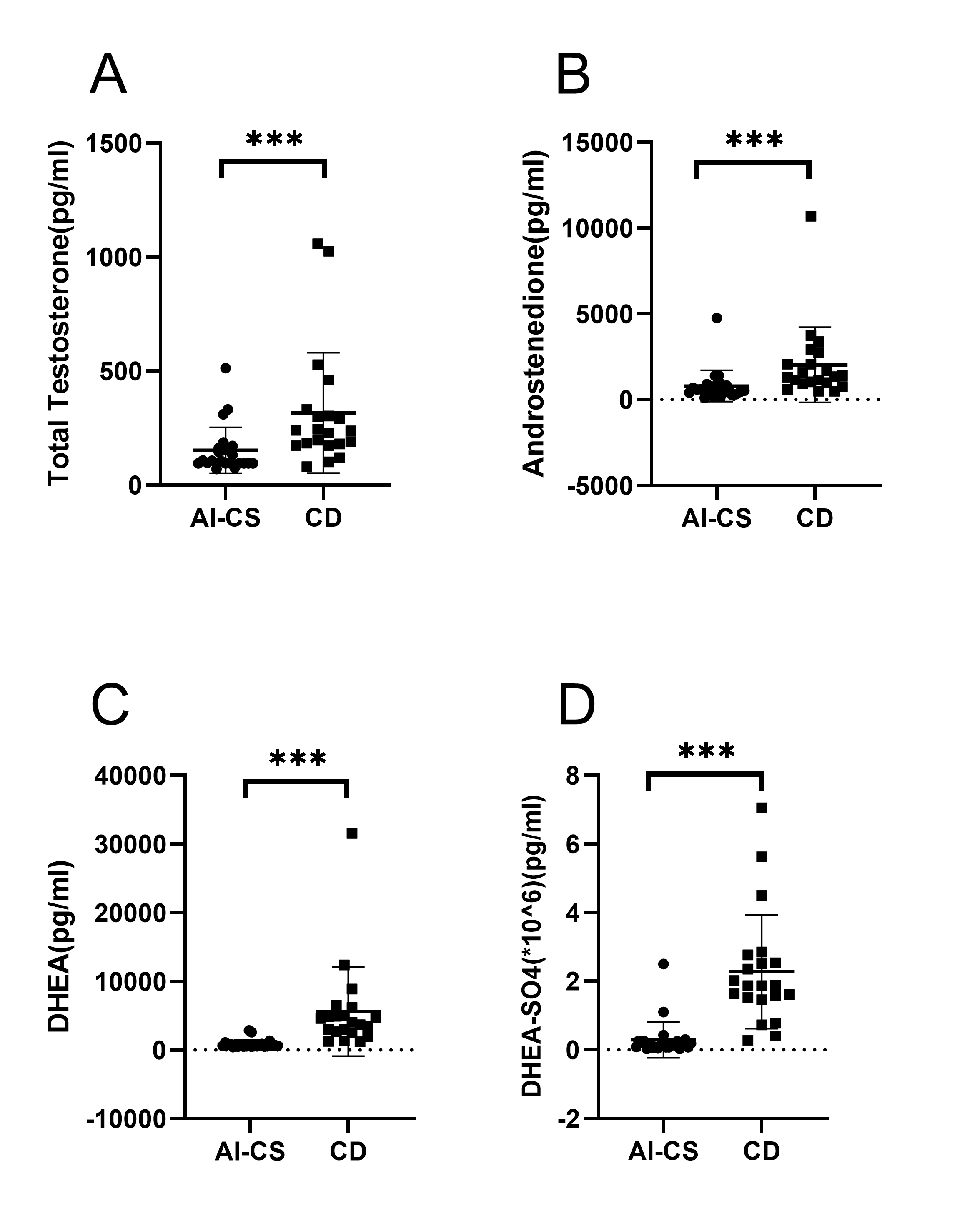

**Supplementary Figure S3. Scatter plots of plasma androgen.** Total testosterone,androstenedione, DHEA and DHEAS values are displayed in (A)-(D), respectively. Comparison between the two groups are shown by brackets, asterisks or text. DHEA, dehydroepiandrosterone; DHEAS, DHEA sulfate; AI-CS, ACTH-independent Cushing’s syndrome; CD, Cushing’s disease. ***, P<0.001.
