## Supplementary Figure S4 for "Impact of Cushing’s syndrome on the hypothalamus-pituitary-gonad axis in women"

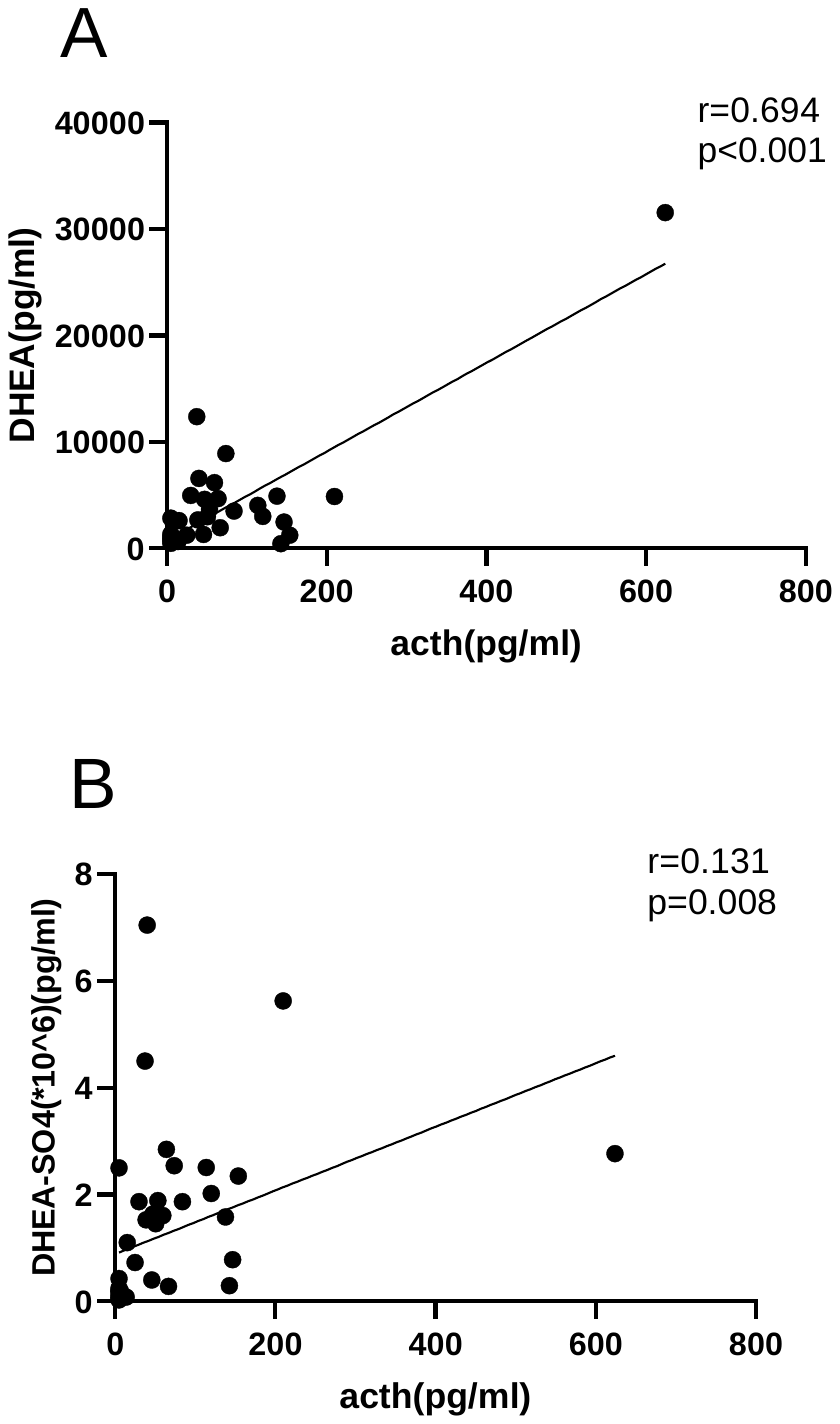

**Supplementary Figure S4. Linear regressions between ACTH and DHEA and DHEAS.** Pearson correlations(r) were calculated squares. DHEA, dehydroepiandrosterone; DHEAS, DHEA sulfate; ACTH, adrenocorticotrophic hormone.
