## Supplementary Figure S5 for "Impact of Cushing’s syndrome on the hypothalamus-pituitary-gonad axis in women"

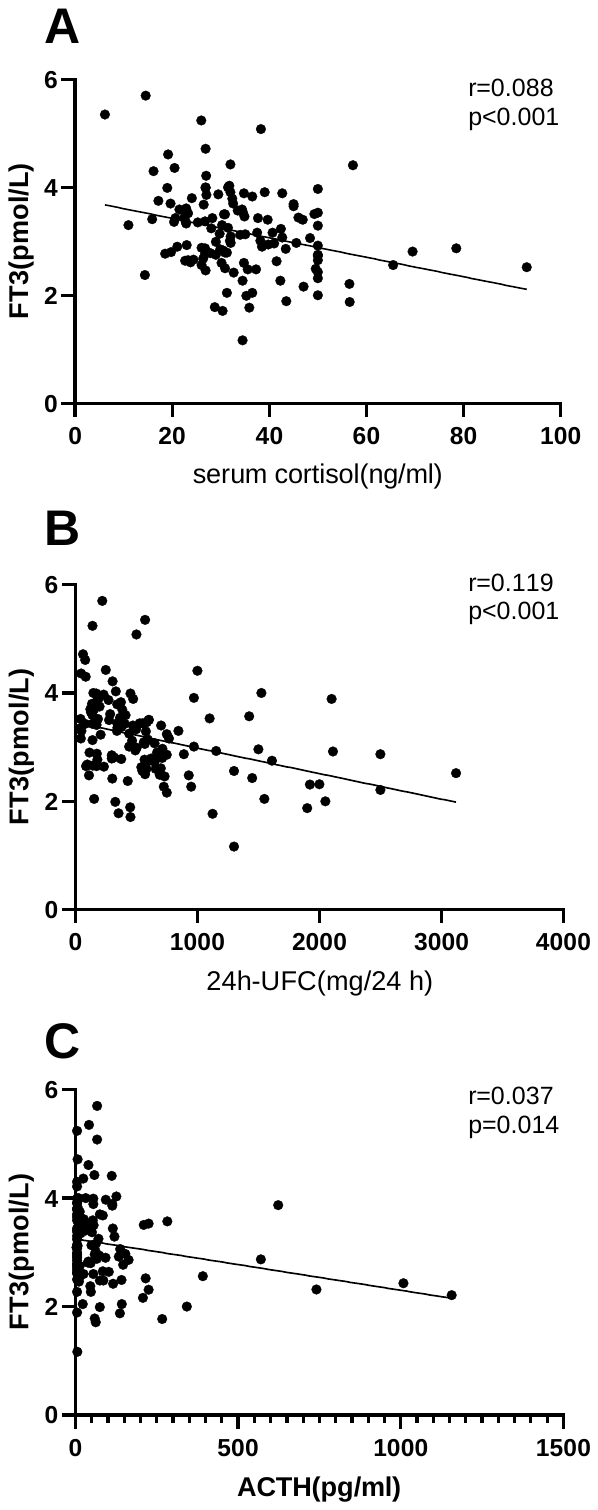

**Supplementary Figure S5. Linear regressions between FT3 and the tensity of hypercortisolism.** Pearson correlations(r) were calculated squares. 24h-UFC, 24h urinary-free cortisol; ACTH, adrenocorticotrophic hormone.
