## Supplementary Table S1 for "Impact of Cushing’s syndrome on the hypothalamus-pituitary-gonad axis in women"

|  | premenopausal cohort | | | | postmenopausal cohort | | | |
| --- | --- | --- | --- | --- | --- | --- | --- | --- |
|  | 24h-UFC | | ACTH | | 24h-UFC | | ACTH | |
|  | r | P value | r | P value | r | P value | r | P value |
| FSH | 0.000 | 0.970 | 0.003 | 0.592 | 0.078 | 0.049 | 0.212 | 0.002 |
| LH | 0.000 | 0.324 | 0.001 | 0.276 | 0.072 | 0.057 | 0.212 | 0.002 |
| TT | 0.100 | 0.001 | 0.256 | <0.001 | 0.063 | 0.07 | 0.197 | 0.003 |

**Supplementary Table S1. Correlation between productive hormones with 24h-UFC and plasma ACTH levels in premenopausal and postmenopausal women.** Pearson correlations (r) were calculated squares. 24h-UFC, 24h urinary-free cortisol; ACTH, adrenocorticotrophic hormone; FSH, Follicle-stimulating hormone; LH, Luteinizing hormone; TT, total testosterone.

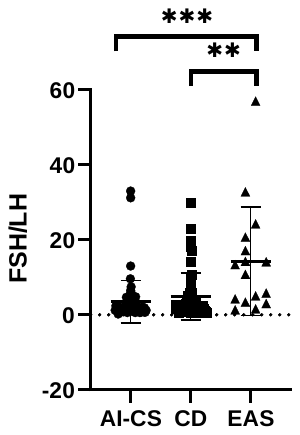

**Supplementary Figure S1. Scatter plots of FSH:LH ratios.** All comparison between the three subgroups are shown by brackets,asterisks or text. FSH, Follicle-stimulating hormone; LH, Luteinizing hormone; AI-CS, ACTH-independent Cushing’s syndrome; CD, Cushing’s disease; EAS, ectopic ACTH syndrome. ***, P<0.001; **, P<0.01.

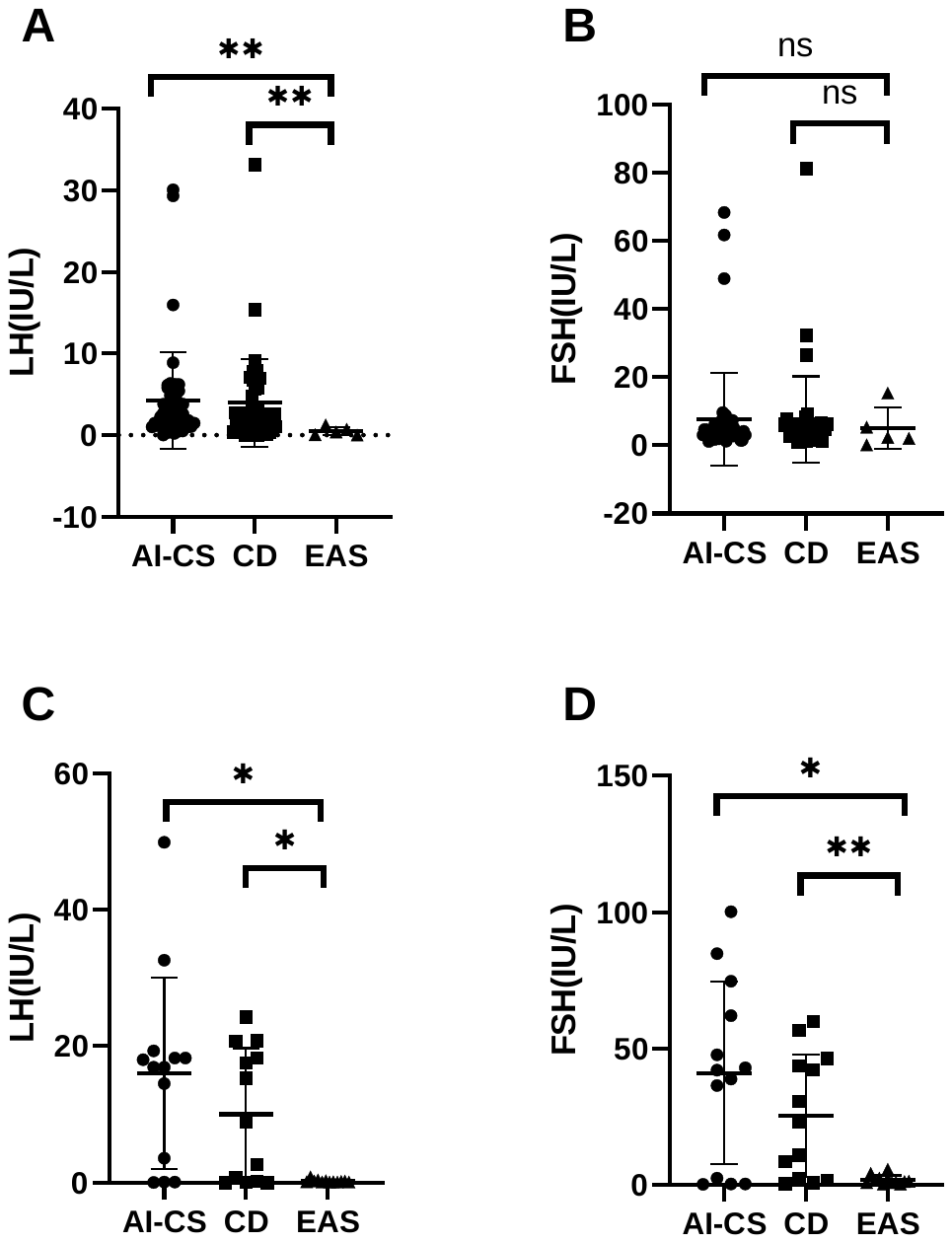

**Supplementary Figure S2. Scatter plots of LH and FSH in women separated by menstruation.** (A)-(B), premenopausal women with CS were included. (C)-(D), postmenopausal women with CS were included. All comparison are shown by brackets,asterisks or text. FSH, Follicle-stimulating hormone; LH, Luteinizing hormone; AI-CS, ACTH-independent Cushing’s syndrome; CD, Cushing’s disease; EAS, ectopic ACTH syndrome. ***, P<0.001; **, P<0.01; *, P<0.05; ns, not significant.

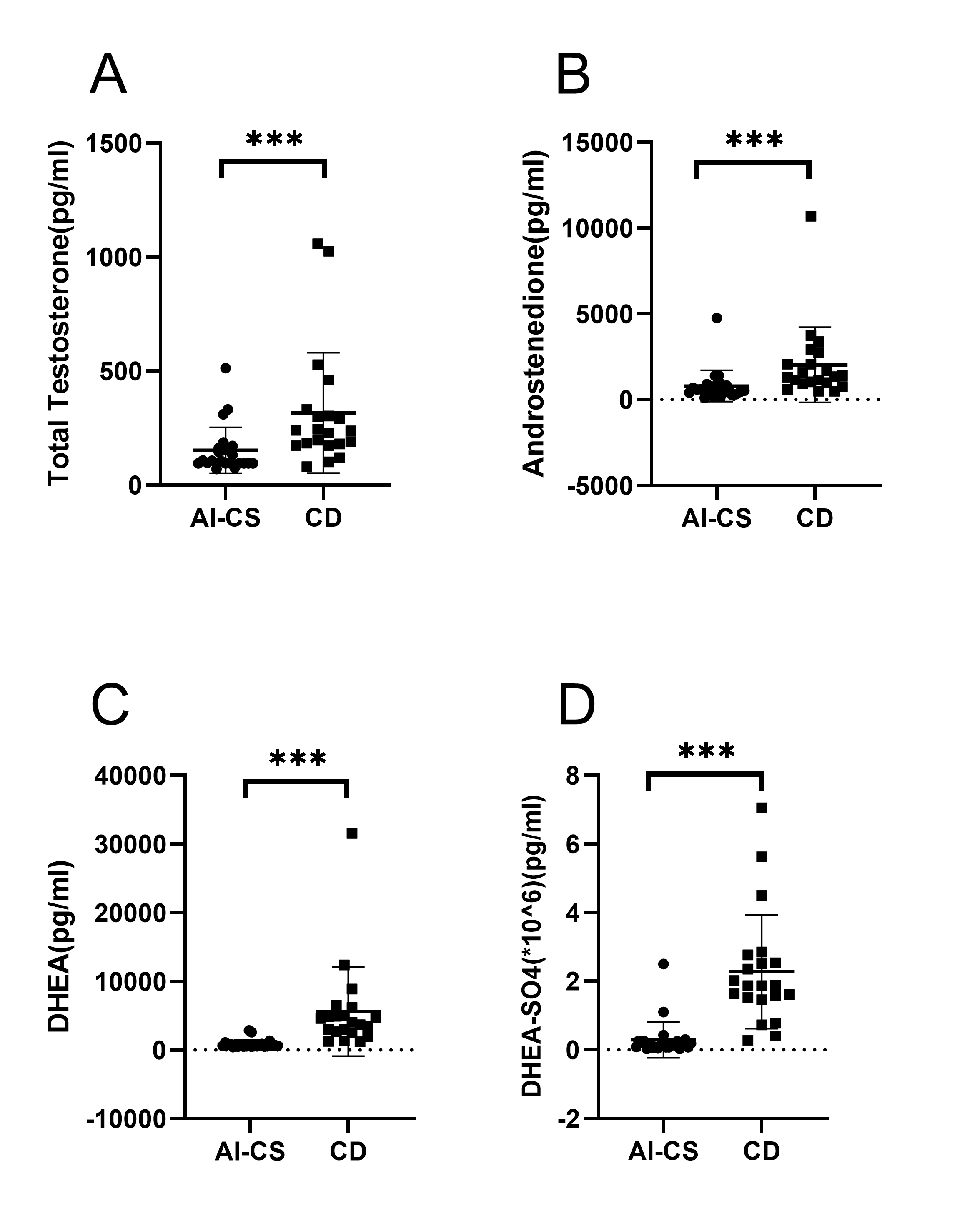

**Supplementary Figure S3. Scatter plots of plasma androgen.** Total testosterone,androstenedione, DHEA and DHEAS values are displayed in (A)-(D), respectively. Comparison between the two groups are shown by brackets, asterisks or text. DHEA, dehydroepiandrosterone; DHEAS, DHEA sulfate; AI-CS, ACTH-independent Cushing’s syndrome; CD, Cushing’s disease. ***, P<0.001.

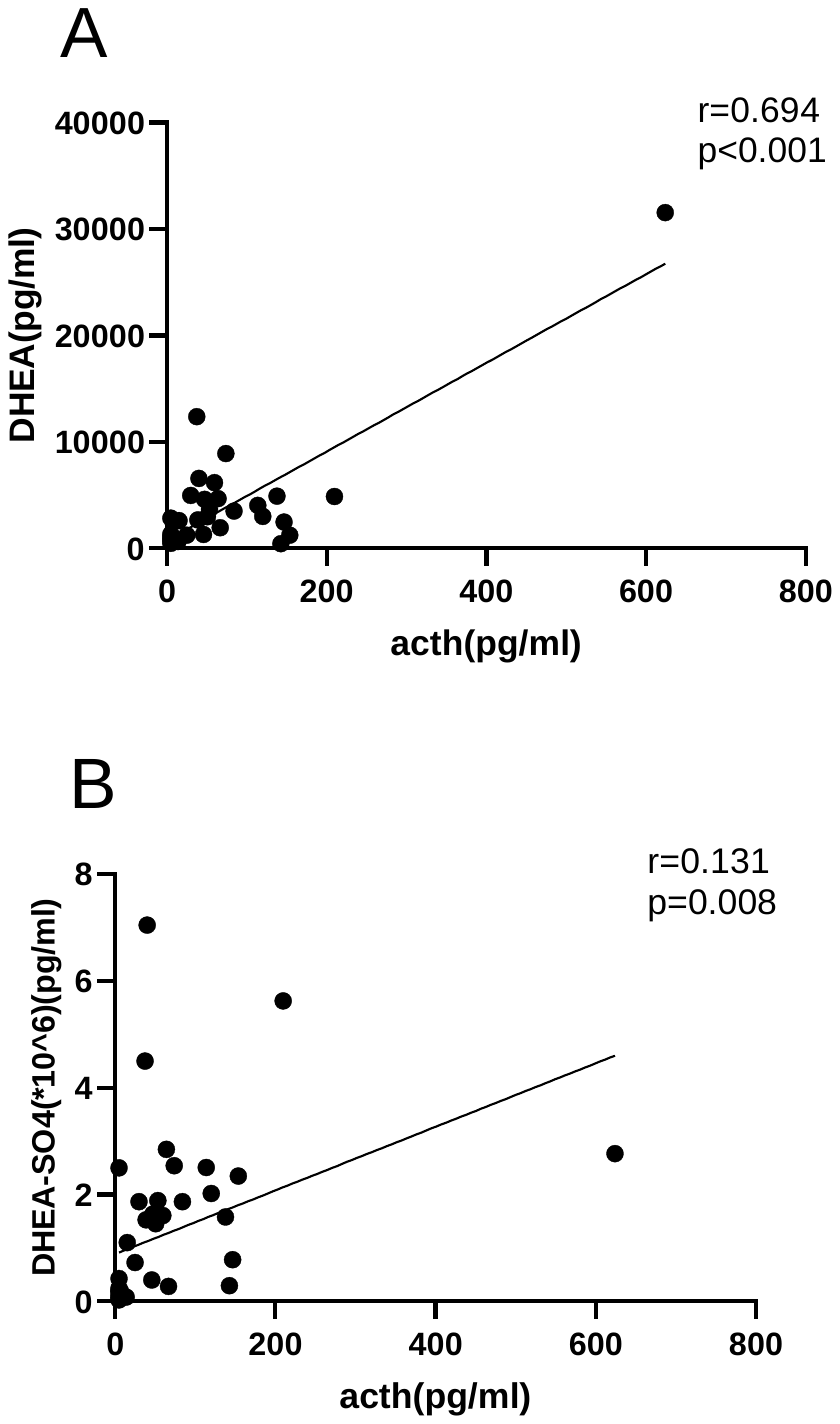

**Supplementary Figure S4. Linear regressions between ACTH and DHEA and DHEAS.** Pearson correlations(r) were calculated squares. DHEA, dehydroepiandrosterone; DHEAS, DHEA sulfate; ACTH, adrenocorticotrophic hormone.

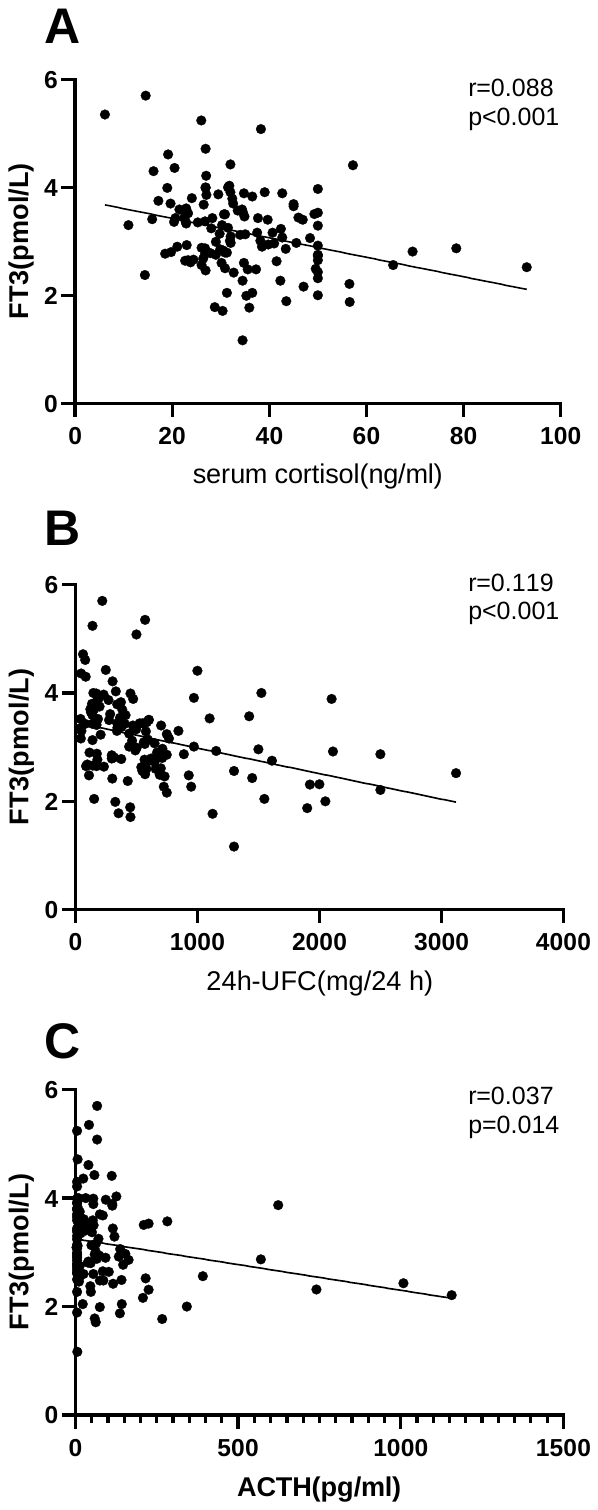

**Supplementary Figure S5. Linear regressions between FT3 and the tensity of hypercortisolism.** Pearson correlations(r) were calculated squares. 24h-UFC, 24h urinary-free cortisol; ACTH, adrenocorticotrophic hormone.
